# Public policy and economic dynamics of COVID-19 spread: a mathematical modeling study

**DOI:** 10.1101/2020.04.13.20062802

**Authors:** Uri Goldsztejn, David Schwartzman, Arye Nehorai

## Abstract

With the COVID-19 pandemic infecting millions of people, large-scale isolation policies have been enacted across the globe. To assess the impact of isolation measures on deaths, hospitalizations, and economic output, we create a mathematical model to simulate the spread of COVID-19, incorporating effects of restrictive measures and segmenting the population based on health risk and economic vulnerability. Policymakers make isolation policy decisions based on current levels of disease spread and economic damage. For 76 weeks in a population of 330 million, we simulate a baseline scenario leaving strong isolation restrictions in place, rapidly reducing isolation restrictions for non-seniors shortly after outbreak containment, and gradually relaxing isolation restrictions for non-seniors. We used 76 weeks as an approximation of the time at which a vaccine will be available. In the baseline scenario, there are 235,724 deaths and the economy shrinks by 34.0%. With a rapid relaxation, a second outbreak takes place, with 525,558 deaths, and the economy shrinks by 32.3%. With a gradual relaxation, there are 262,917 deaths, and the economy shrinks by 29.8%. We also show that hospitalizations, deaths, and economic output are quite sensitive to disease spread by asymptomatic people. Strict restrictions on seniors with very gradual lifting of isolation for non-seniors results in a limited number of deaths and lesser economic damage. Therefore, we recommend this strategy and measures that reduce non-isolated disease spread to control the pandemic while making isolation economically viable.

## Introduction

As of May 1st, 2020, the number of confirmed cases of coronavirus disease 2019 (COVID-19) worldwide stands at over 3,240,000, and at least 230,000 individuals have died from this disease since the first reports of a pneumonia of unknown etiology in December of 2019 (1; 2; 3). As COVID-19 expands to new territories, after a latent phase with few new reported infections daily, the virus spreads rapidly and local outbreaks begin. The virus’s containment is complicated by its high transmissibility, relatively low case fatality rate, and lengthy asymptomatic infectious period (4). Currently, the most successful measure to prevent the rapid expansion of COVID-19 has been to implement isolation policies to restrain travel and physical interactions between individuals (5; 6). Extreme isolation policies have devastating consequences for the global economy. Isolation policies may be an effective short-term measure, but indefinite isolation until a vaccine becomes available would prevent billions of individuals worldwide from receiving an income and could particularly devastate countries with weaker economies. This economic harm could increase mortality given the correlation between mortality and income (7; 8).

Computational models that simulate the expansion of the disease can guide public policy makers in mitigating the detrimental outcomes of the outbreak (9). Existing epidemiological models estimate the number of unknown infected individuals, forecast the number of patients requiring intensive care treatment, predict the death toll, and evaluate the outcomes of different policies.

In compartmental modeling, different disease stages are modeled as compartments, and transitions between compartments are modeled by a system of differential equations (10). Compartmental models provide useful insight into the mechanisms governing the spread of the disease and help evaluate public policies (11; 12; 13). Some studies focus on how isolation restrictions affect disease spread (6; 13; 14; 15; 16). Time series approaches use available data to statistically forecast the evolution of the disease (17; 20; 21). While time series models have precise results, they do not illuminate the dynamics of the disease spread. To accurately predict the spread of disease and evaluate consequences beyond the infectious disease itself, models must take into account how mitigation measures might impact the economy (22; 23; 24). Existing COVID-19 models usually do not integrate the economic aspects of public policy aimed at controlling the spread of the disease. Few models incorporating public policy responses take into account both the distinctions in risk types within the population and the differential economic impacts of isolation policies. Glover et al. (25) build this kind of model with only the non-seniors working and focus on differences in policy preferences between different age groups in the economy. Acemoglu et al. (26) also study the gains of targeting isolation policies differently based on age risk as compared to uniform isolation policies.

We have developed a predictive model for COVID-19 that considers, for the first time, the intercoupled effects of different isolation policies on both economic and health outcomes. We separate the population into four groups, according to age and the economic impact of remaining isolated for an extended period of time. Upon infection, senior individuals present much larger comorbidity and mortality rates than non-senior individuals (19; 18). Our model includes, among other compartments, compartments that represent isolated, infected, hospitalized, recovered and dead individuals. For simplicity, our infected compartment contains all infected individuals who are not hospitalized, including both those that are showing symptoms and those who are not showing symptoms and who may not know that they have the disease.

Also, generalized country-wide isolation policies completely suppress the productivity of some workers while having minimal implications for others. By modeling our population as having different health risk groups, economic risks from isolation, and productivity levels, we can make more refined predictions and are able to provide more targeted policy recommendations than existing methods.

We model public policies that balance the number of total deaths and damage to economic productivity because public policymakers are likely to incorporate economic effects and how citizens perceive isolation restrictions within their policies. These policies greatly affect the state of the economy, which can influence the health of citizens. In addition, we model improvements in medical knowledge about a recently discovered disease as decreasing the hospitalization fatality rate over time, i.e., the fraction of patients that require hospitalization and eventually die, motivated by evidence of improved treatments and mortality outcomes for COVID-19.

Our model captures the main dynamics of disease and policy responses to rigorously model the trade-offs between economic damage and health outcomes. Based on the available evidence, we identify the main aspects of the pandemic to be the rate of disease spread, the differences in outcomes by age, the improvements in health outcomes for diseased patients over time, and economic damage from the pandemic. While we use government policy as the mechanism for placing individuals in an out of isolation, individuals can also choose to self-isolate or stop isolating based on economic conditions and disease spread. Our model applies equally whether people are isolated by the government or whether they isolate themselves. Our simulations also clarify differences between short-term and long-term outcomes from a pandemic, both for health and economic outcomes, and the consequences of prioritizing short-term outcomes over long-term outcomes. There may be short-term economic gains from certain government or individual decisions to stop isolating, but they may lead to future economic losses. Similarly, while the current level of disease spread might be low, decisions to stop isolation policies in response to small amounts of disease can lead to many future deaths.

Our simulations show that over 76 weeks and in a population of 330 million individuals, which imitates the population of the USA: (i) if the current initial stringent isolation policies are kept in place, where 85% of the non-seniors and 95% of the seniors are isolated, the outbreak is controlled about 300 days from its start and there are 207,906 deaths. Additionally, the economy shrinks by 34.0% with respect to the pre-pandemic state. (ii) If isolation restrictions for non-seniors are rapidly lifted shortly after outbreak containment, a second outbreak takes place. This prolongs the duration of the pandemic and necessitates the re-institution of isolation restrictions. In this scenario, there are 525,558 deaths and while the economy expands shortly after lifting isolation restrictions, it shrinks rapidly after reinforcing isolation restrictions. Overall, the economy shrinks by 32.3% by the end of the simulation. (iii) Lastly, with a very gradual relaxation of isolation restrictions for the non-seniors, the number of deaths is slightly higher than the number in the baseline scenario and the economy recovers slowly but steadily. Overall, the economy shrinks by 29.8% with respect to the pre-pandemic state.

## Results

### Model overview

We develop an expanded SEIR model, incorporating different levels of disease risk and economic damage from isolation, as well as improved disease knowledge over time. We present the health and economic dynamics of our model.

#### Epidemiological model and dynamics

To model the spread of COVID-19, we design a compartmental model with ten interconnected compartments. Each compartment is subdivided into two parts, one containing the senior population and the other containing the rest of the population.

The transition rates of the compartments are illustrated in Fig. 1, and the variables used are listed in Table 1. Susceptible individuals get exposed to COVID-19 at a rate proportional to the density of infected individuals and hospitalized individuals. Exposed individuals then become infected, where we define infected patients as all non-hospitalized infected individuals. These individuals can either recover from their current state or develop serious symptoms and become hospitalized. The fraction of infected individuals that become hospitalized follows a Bernoulli distribution. In other words, the infected individuals become hospitalized with probability *p* and recover without requiring hospitalization with probability *q* = 1 − *p*. The probability of requiring hospitalization is lower for the general population and higher for the senior population.

**Figure 1:**
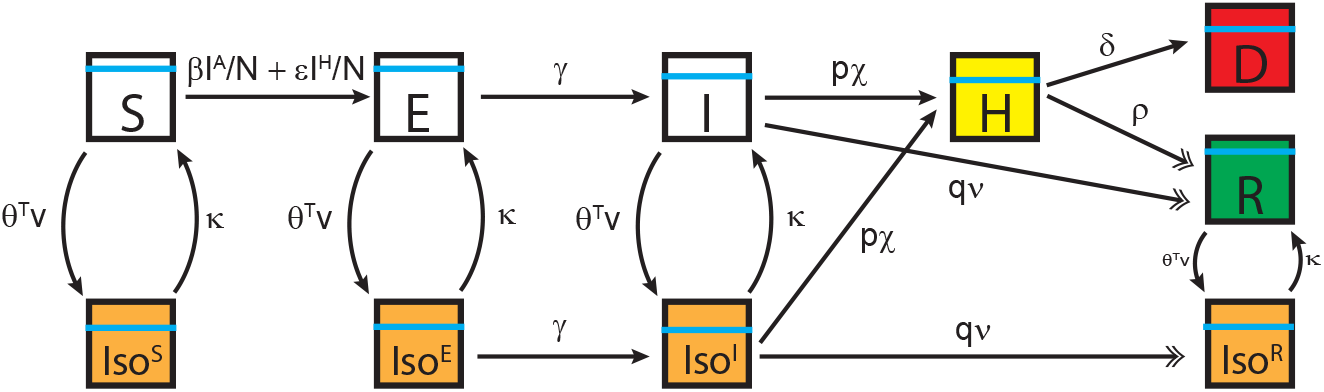
Compartmental model. Each box represents an individual compartment and the arrows represent transitions between the compartments. The individuals in every compartment are divided by age, as represented by the cyan bars, and by their productivity, as represented by the background color of the boxes. White boxes represent normal productivity, dead individuals have no productivity, which is represented in red, isolated individuals have decreased productivity, which is represented in yellow, and hospitalized individuals have no productivity and incur treatment costs, which are also represented in red. As individuals recover, they receive a small boost in productivity due to their acquired immunity if they are in isolation, which is represented in green.

**Table 1:**
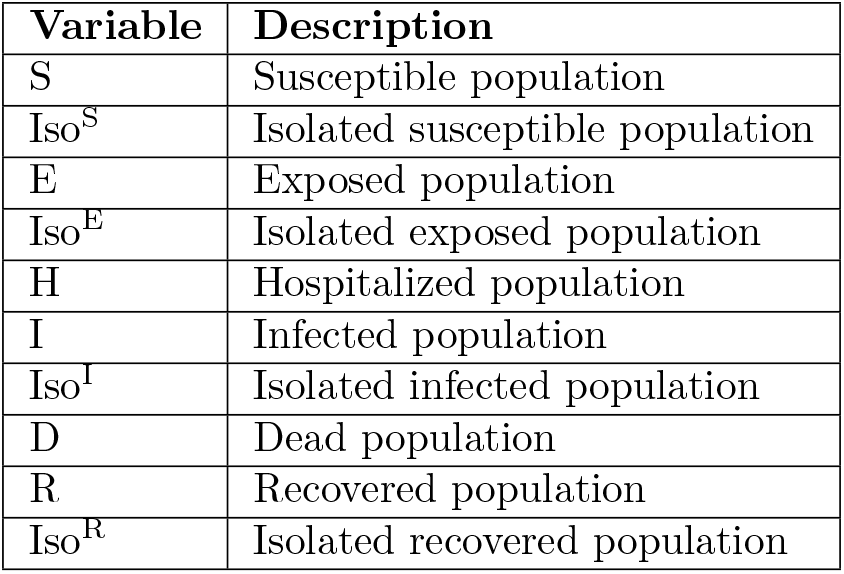
Variables used in the compartmental model.

**Table 2:** Summary of predicted results from the different scenarios.

| Scenario | Baseline | Sudden release | Gradual Release |
| --- | --- | --- | --- |
| Peak number of hospitalizations | 183,984 | 2,258,337 | 183,984 |
| Non-senior deaths | 21,230 | 154,230 | 26,399 |
| Senior deaths | 214,494 | 371,328 | 236,518 |
| Net change in productivity | -34.0% | -32.3% | -29.8% |

The hospitalized individuals either die or recover at appropriate rates. These rates depend on whether the hospitalized population is above hospital capacity strain and saturation points. The hospitalization fatality rate decreases over time to account for better knowledge by the healthcare system.

Individuals who are not hospitalized enter or leave isolation compartments according to public policy or their own individual decisions. We model the transition rates into and out of isolation through *θ*′ · *υ* and *κ*, respectively. Where v is a vector containing the number of reported infections, deaths, and the current state of the economy, and *θ* is a weight vector that reflects the relative influence of the values in υ to drive the population into isolation. Lastly, *κ* is the rate at which individuals transition out of the isolation states. Throughout our simulations, *θ*′ · *υ* ≥ 0 and *κ* ≥ 0. When *θ* = *κ* = 0, then the fraction of the population in isolation remains constant

The transition rates between the compartments, which are represented with Greek letters, are derived from published data about the progression rate of COVID-19 and are described in the supplementary material (S1 Appendix and S1 Table).

#### Economic Model and Dynamics

In our model, there are occupations more affected by isolation and occupations less affected by isolation. In our simulation, we use two types of workers: those who have higher productivity in isolation and those who have lower productivity in isolation. The total economic output is the sum of the outputs of the individuals in each group minus the costs of treating the infected. For the longterm economic output beyond the pandemic period, deaths impose a penalty on future economic output, dependent on the discount rate *r*.

The productivity of each labor group is the sum of the productivity of all the individuals in that labor group. The cost of treating the infected is the number of individuals of each risk type times the cost of treating each risk type. Different workers of the same labor type might be under different isolation restrictions based on their age or risk type. In our simulation, there are four levels of productivity for four types of workers: older workers who have higher ability to work in an isolated state (*y*_S,H_), senior workers who have lower ability to work in an isolated state (*y*_S,L_), non-senior workers who have higher ability to work in an isolated state (*y*_G,H_), and non-senior workers who have lower ability to work in an isolated state (*y*_C,L_). The workers who have a lower ability to work in an isolated state lose a higher percentage of their productivity (represented by *ψ*) than workers who have a higher ability to work in an isolated state (represented by *ϕ*). In our simulation, we use college education as a proxy for ability to work in an isolated state, as found by Mongey et al. (37).

The dynamics of output in this model depend on the rate at which workers in different productivity and health groups move in and out of isolation and the rates with which their health states change. A complete mathematical description of the model (S1 Appendix) and the values used for the transfer rates (S1 Table) appear in the supplementary material. Additionally, a full simulation, showing the evolution of the number of individuals in every compartment is shown in Supplementary Fig. S1.

The epidemiological model and the economic model are coupled. The epidemiological model is used to calculate the spread of COVID-19 together with the number of hospitalizations and deaths. These values, in turn, impact the imposition of isolation policies, which decrease the economic output. The decreased economic output, calculated through the economic model, can lead to calls for less strict isolation policies. As a result, the interaction of both models describes many aspects of the COVID-19 pandemic.

Using a population of 330 million, which mimics the population of the United States, we simulate the evolution of public policy given the preferences of the policymakers over 76 weeks. In each scenario, to imitate strict isolation policies, we start with 85% of non-seniors and 95% of seniors in isolation. Public policy and individual choices shift the population in and out of isolation. We keep the non-isolated population above zero because some jobs that could potentially spread disease continue largely unchanged, since they are deemed essential in a pandemic economy. For this reason, there is not much room for further strengthening isolation for non-seniors. The initial conditions are meant to reflect a relatively early stage of an epidemic, where disease has become widespread enough to make traditional preventative and contract tracing measures inadequate, but where most of the population has not experienced the disease. The initial conditions are detailed in the supplementary material (Supplementary Table S3).

### Baseline scenario

In the baseline scenario, we keep the current isolation status constant. (*θ* = 0, *κ* = 0). That is, the initial population in isolation remains isolated for the entire duration of the simulation (Supplementary Table S2). The number of non-seniors and seniors infected but not hospitalized is shown in Fig. 2A and 2D, respectively. Leaving isolation measures in place results in 235,724 total deaths. The peak number of hospitalized is 183,984, which does not strain hospital capacity, as shown in Fig. 2C. Total deaths first continue to grow as more non-isolated individuals get infected, but then level off. There are 21,230 deaths among non-seniors and 214,494 among seniors, as shown in Fig. 2B and Fig. 2E, respectively. Since both non-seniors and seniors have strict isolation restrictions imposed on them, most of the deaths occur in the senior population, which is more vulnerable to the disease.

**Figure 2:**
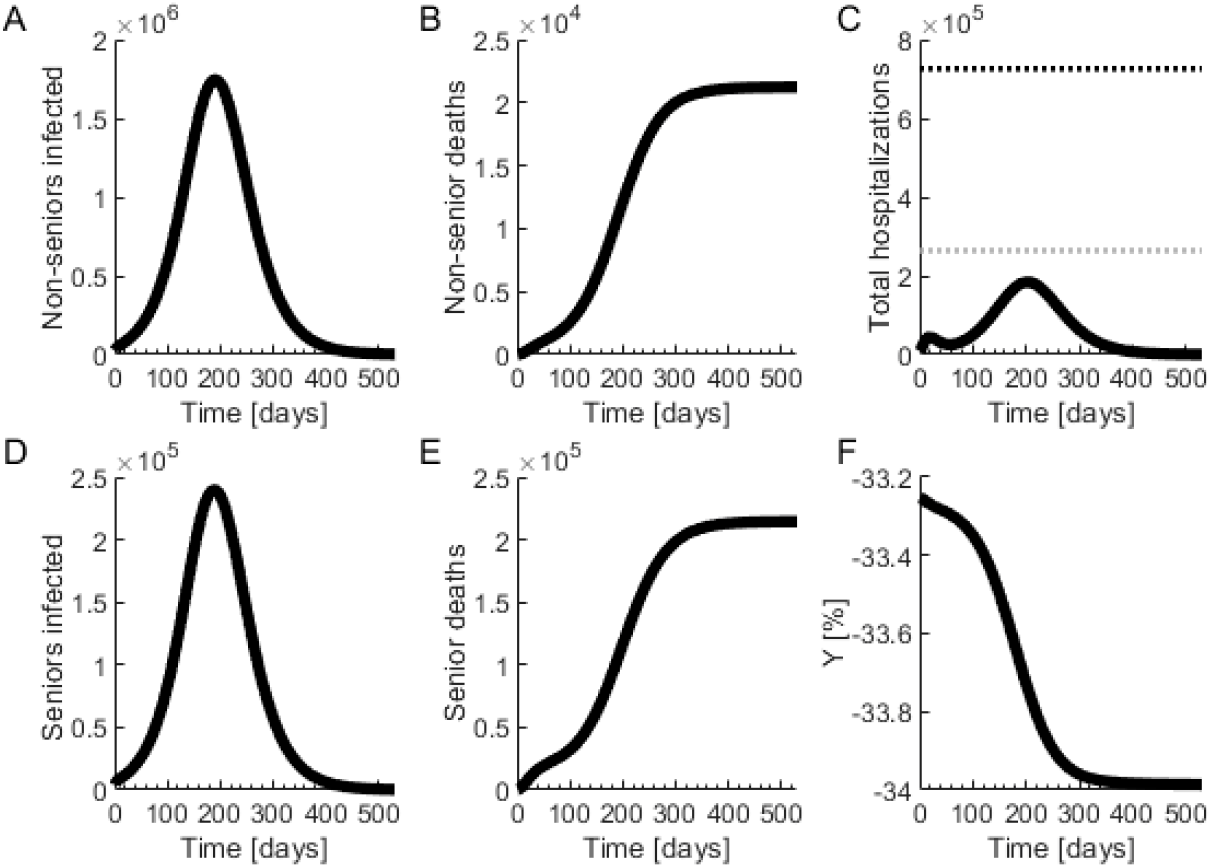
Outcomes in the baseline scenario. (**A**) Number of infected non-senior individuals. (**B**) Number of non-senior deaths. (**C**) Total hospitalizations. The gray dotted line marks the hospital capacity strain level and the black dotted line marks the saturation level. (**D**) Number of infected senior individuals. (**E**) Number of senior deaths. (**F**) Change in economic output compared to the pre-pandemic state.

The economy in the baseline scenario, shown in Fig. 2F, shrinks slightly from its initial restricted levels, as some people are hospitalized and die. This is costly for the economy, and there is no increase in productivity from others as productivity is still limited by isolation restrictions. Given that the initial restrictions are kept throughout the entire simulation time and have a strong negative impact on economic output, the economic productivity in this scenario is quite low for a prolonged period of time.

### Sudden release of the population after control of outbreak, but before complete eradication

In this scenario, we maintain the isolation policy detailed in the baseline scenario, and then release the only the non-senior isolated population at a rate of 3% daily. This release rate is simulated by modifying the value of *κ*. We release the population when the number of infected individuals is decreasing and becomes less than 0.2 % of the total population, which in our model takes place in week 41. If strict isolation measures bring the disease largely under control earlier, then the population can be released at an earlier time. If the hospitalized population reaches 85% of the hospital capacity saturation point, isolation restrictions are restored. Individuals transit to the isolation compartments by setting *κ* to zero, and increasing *θ* (Supplementary Table S2). As can be seen in Fig. 3A and Fig. 3D, a large second outbreak soon takes place after the release of the population, which necessitates the re-enforcement of isolation policies. After releasing the population, the number of hospitalizations increases until saturating the health care capacity about seven weeks after the release. This happens even after isolation restrictions are re-imposed, as shown in Fig. 3C. Owing to the sudden release, the peak number of hospitalized is 2,258,337, which saturates the hospital capacity and increases the hospitalization fatality rate. There are 525,558 total deaths, with 154,230 deaths among non-seniors, as shown in Fig. 3B, and 371,328 among seniors, as shown in Fig. 3E.

**Figure 3:**
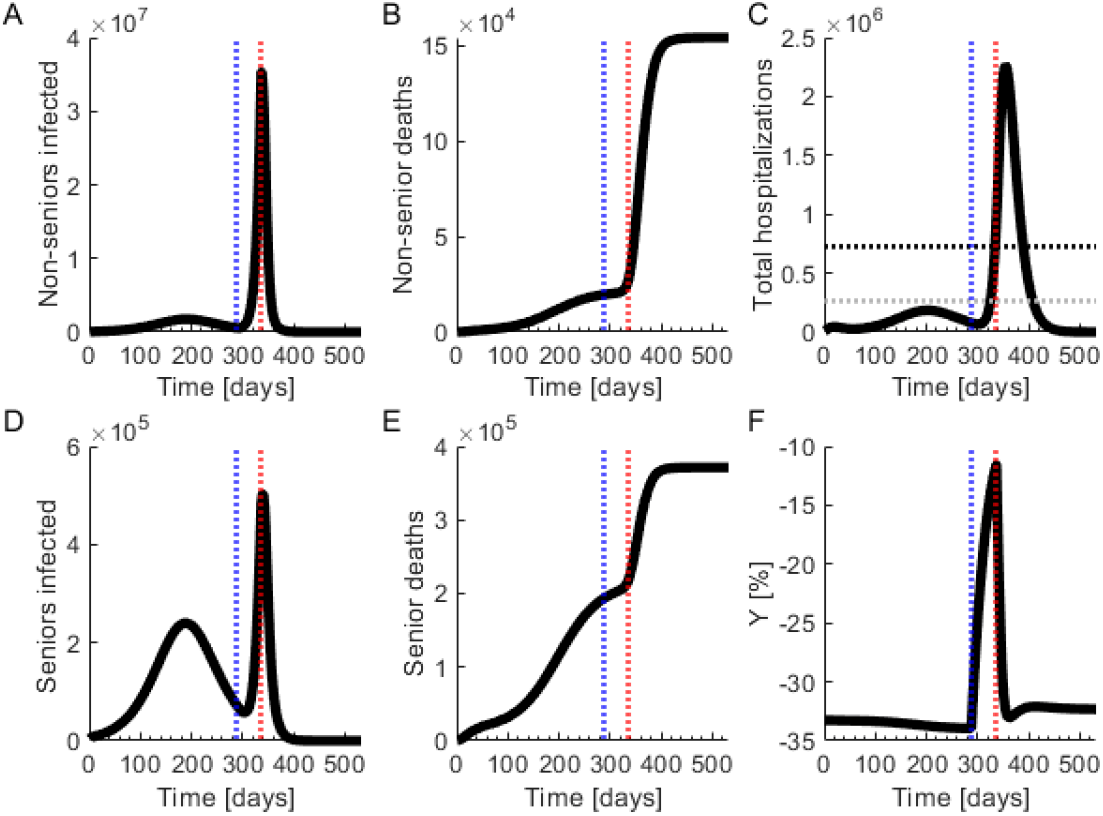
Sudden release of isolation measures after outbreak containment. Completely relaxing the isolation measures after the outbreak is controlled but before completely eradicating the virus leads to a second outbreak. Isolation measures are re-enforced after the second outbreak takes place and hospitals become saturated. (**A**) Number of infected non-seniors. (**B**) Number of non-senior deaths. (**C**) Total hospitalizations. The gray dotted line marks the hospital capacity strain level and the black dotted line marks the saturation level. (**D**) Number of infected seniors. (**E**) Number of senior deaths. (**F**) Change in economic output compared to the pre-pandemic state. The vertical blue dotted lines mark the time at which isolation policies are lifted and the vertical red lines mark the time at which they are re-enforced.

Figure 3A shows a decreasing number of infections before the second outbreak. During this initial period, the very low number of newly infected and deaths may suggest that COVID-19 has been successfully mitigated, and the pressure to resume normal activities may grow. The economy is strongly boosted shortly after strict restrictions are removed. However, due to the large initial fraction of non-hospitalized infected individuals, a sudden relaxation of isolation restrictions after the disease is largely controlled leads to a second outbreak, with increased deaths and infections. The economic dynamics of this scenario are shown in Fig. 3F. After a short-term economic boost, the second outbreak damages economic productivity later, as shown in Fig. 3F.

### Progressive restart of the economy before the pandemic is over

In this scenario, we investigate a gradual relaxation of isolation restrictions. Various studies have shown that the case fatality rate and the hospitalization fatality rate for the senior population are larger than that of the non-senior population (28; 29). The marked difference in mortality according to age suggests that enforcing an extremely strict isolation policy for the senior population while allowing the general population to slowly resume normal activities could potentially balance disease spread and economic damage.

In this scenario, as in the sudden release scenario, policymakers loosen isolation restrictions when the number of infected individuals decreases below 0.2 % of the total population, releasing the non-senior isolated population but at a rate of 0.1% daily (Supplementary Table S2). This gradual relaxation does not lead to a second outbreak, as can be seen from the number of infected individuals in Fig. 4A and Fig. 4D. In this state, the peak number of hospitalized is 183,984, which strains but does not saturate the health care system, as shown in Fig. 3C. There are 26,399 non-senior deaths and 236,518 senior deaths, as shown in Fig. 4B and Fig. 4E, respectively. In this scenario, there are 27,193 more deaths than in the baseline scenario and a 29.8% decrease in economic productivity, with output trending upwards as more individuals are released from isolation. As we see in Fig. 4F, relaxing restrictions on seniors as well does not improve economic productivity and results in substantially more deaths, as shown in the red traces of Fig. 4. Additionally, if the rate at which the general population abandons isolation measures increases rapidly, a second outbreak could take place, as this brings us closer to the sudden release scenario. Strict enforcement of isolation restrictions for seniors is vital, as without strict enforcement for senior individuals, the number of senior deaths increases rapidly (Fig. 4E) once the non-seniors start getting infected at high rates (Fig. 4A).

**Figure 4:**
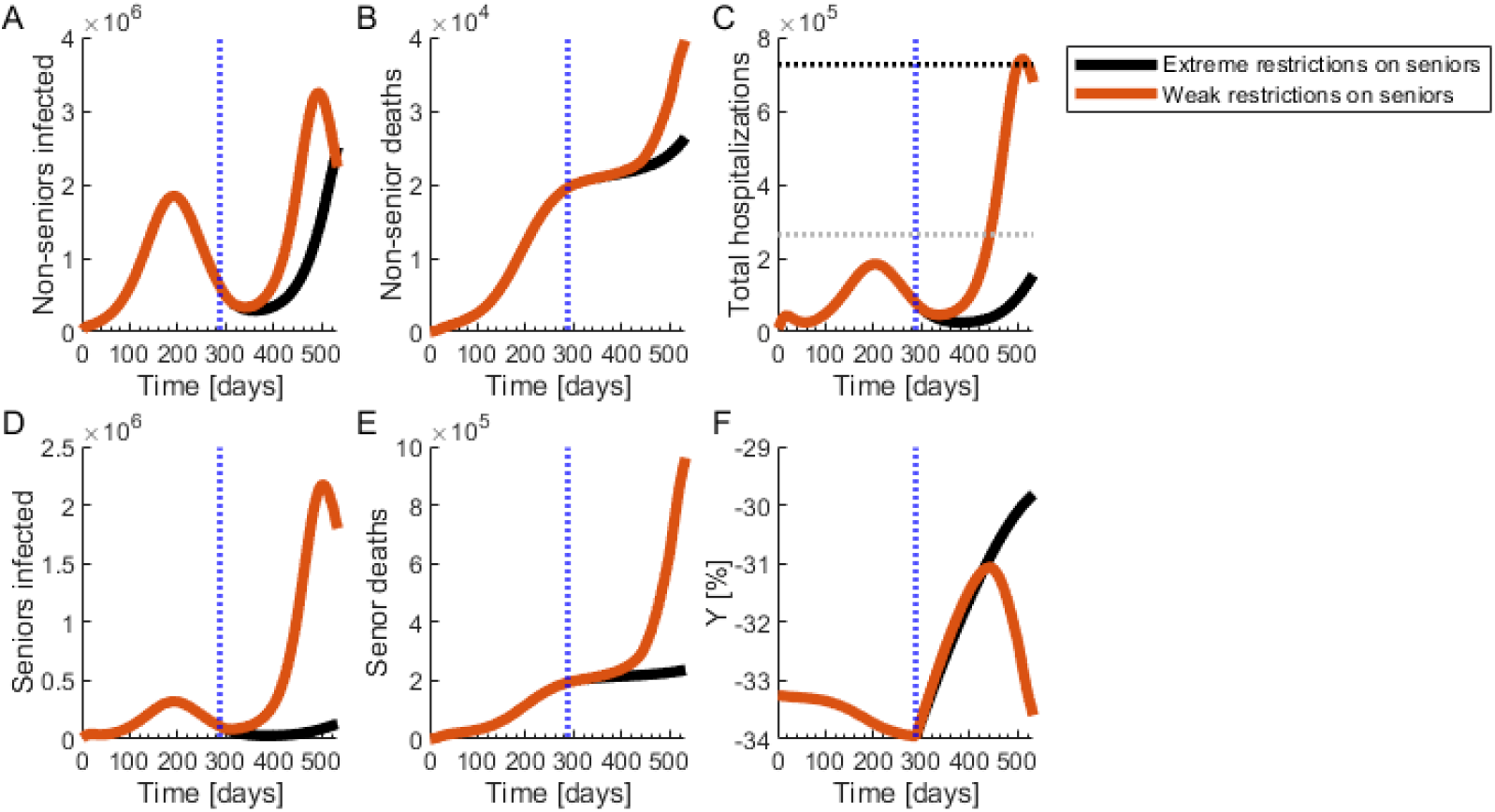
Progressive release of the non-senior population with extremely strict isolation measures for the seniors. (**A**) Number of infected non-seniors. (**B**) Non-senior deaths. (**C**) Total hospitalizations. The gray dotted line marks the hospital capacity strain level and the black dotted line marks the saturation level. (**D**) Number of infected seniors. (**E**) Number of senior deaths. (**F**) Change in economic output compared to the pre-pandemic state. (**A-F**) The black curve represents the scenario where the isolation policies for the seniors are extreme. The red curve represents the scenario where the policy is weakly implemented and the isolation measures for the senior population are not extreme. The vertical blue dotted lines mark the time at which isolation policies are lifted.

Economic output is higher in the gradual isolation relaxation scenario with strict enforcement of isolation for seniors (see Fig. 5D). However, while controlled disease spread and the highest economic output occurs in this scenario, the pandemic still does not disappear, meaning that relaxing isolation policies for the senior population at any time leads to an extremely large number of hospitalized and dead individuals, as shown in Fig. 4D. Additionally, a change to a more rapid relaxation policy in the future will also lead to a second outbreak. Therefore, although this scenario can lead to an acceptable economic situation with a controlled number of hospitalizations and deaths, it is not a viable longterm solution. Any isolation policy that does not eliminate the disease can serve only as bridge to a global vaccination campaign.

**Figure 5:**
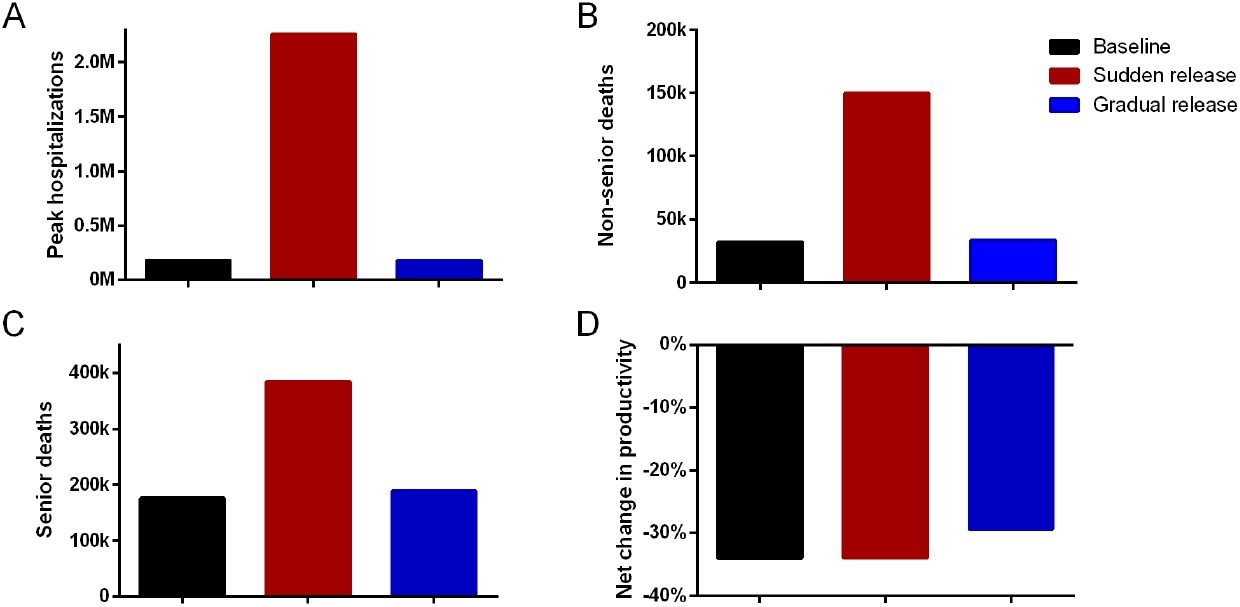
Comparison of all scenarios. (**A**) Peak number of hospitalizations. (**B**) Non-senior deaths. (**C**) Senior deaths. (**D**) Net change in productivity.

As Fig. 5 shows, health outcomes in the sudden release scenario are much worse, with many more hospitalizations and more deaths. Despite a temporary bump in economic output when isolation restrictions are loosened, by the end of the 76 week period, economic output in the sudden release scenario is similar to output in the baseline scenario. The gradual release scenario does not saturate the hospital system, with the same peak number of hospitalizations as in the baseline scenario, and there are 27,193 additional deaths as compared to the baseline scenario, which has 11.5% more deaths. The gradual release scenario has a higher economic output than the baseline scenario and the sudden release scenario, with a −29.8% change in productivity since the pre-pandemic state, as compared to a −34.0% change in the baseline scenario and a −32.3% change in the sudden release scenario.

### Effect of public policy on controlling the spread of COVID-19

We perform a sensitivity analysis on the parameters that are sensitive to public policy to evaluate the outcomes of different policy decisions. We vary each parameter which can be affected by public policy, while keeping the rest of the model parameters constant, and examine the effect of adjusting each parameter on hospitalizations, deaths, and economic output. First, from Fig. 6A-C, we observe a large decrease in hospitalizations and deaths, and a large increase in economic productivity, as contagiousness of infected non-hospitalized patients (*β*) is controlled. We also observe that the total number of hospitalizations and deaths decrease slightly with the control of contagiousness of hospitalized patients (*ϵ*), while economic productivity grows slightly. This observation suggests the importance of public health measures that slow disease spread among non-isolated individuals that do not know their disease status.

**Figure 6:**
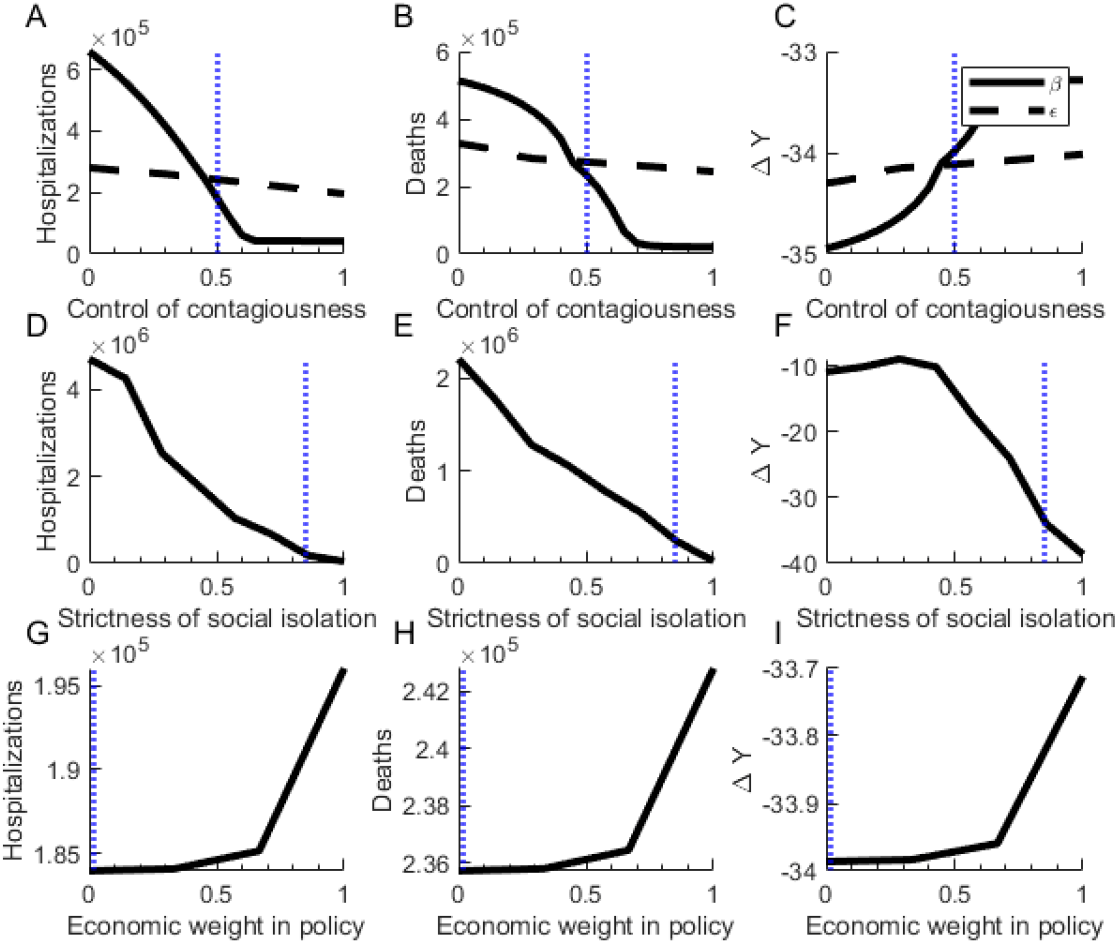
Sensitivity analysis. **A-C**. Total hospitalizations, deaths, and net change in economic productivity for one and a half years depending on the control of contagiousness of asymptomatic (*β*) and hospitalized (*ϵ*) individuals. **D-F**. Total hospitalizations, deaths, and net change in economic productivity for one and a half years depending on the strictness of enforcement of isolation policies. **G-I**. Total hospitalizations, deaths, and net change in economic productivity for one and a half years depending on the relative importance of the economic situation in the public policies. The blue dotted vertical lines mark the values used in the baseline scenario.

Secondly, from Fig. 6D-F, we observe that as the strictness of isolation measures increase, the number of deaths and hospitalizations decrease, while economic output is first flat, and then at certain level of isolation restrictions, begins to sharply decrease. Meanwhile, when increasing the strictness of isolation restrictions, the number of hospitalizations and deaths decreases. Strict isolation measures target the number of people sent in and out of isolation, holding other parameters constant.

Maximum number of simultaneous hospitalizations during the pandemic, deaths in the non-senior and senior populations, and net change in productivity from the pre-pandemic state to a year and a half from the beginning of initial conditions of the model. In the baseline scenario, the restrictions do not change. In the sudden release scenario, after the disease is mostly brought under control, restrictions for non-seniors are suddenly relaxed. In the gradual release scenario, after the disease is mostly brought under control, restrictions for non-seniors are gradually relaxed.

Finally, we see in Fig. 6G-I that as policymakers put more emphasis on economic productivity, there are increases in hospitalizations and deaths, and there is a small increase in economic productivity. We see that if policymakers value short-term economic productivity highly relative to disease spread, there is a danger of isolation policies leading to larger death tolls.

## Discussion

We present a model of COVID-19 in a population with different risk groups and different levels of economic vulnerability to public policy aimed at preventing disease spread. We focus our model on a situation where disease is ongoing and isolation measures are already in place. By modeling our population with different risk groups and economic groups, we are able to produce specific policy suggestions that allow for a finer targeting of the spread of COVID-19. The seniors are most vulnerable to COVID-19 and can be protected from the disease without crippling the economy. We suggest stringent isolation policies for higher-risk groups. Our simulations show that in order to protect the most vulnerable, any relaxation of isolation restrictions must be gradual, even for the less vulnerable.

Beyond the short-term, isolation policy does not need to be as strict for lower-risk groups. Isolation restrictions can lessen over time for lower-risk people while still minimizing deaths. Lessening isolation restrictions allows the economy to recover to −29.8% of pre-restriction productivity, as opposed to −34% when isolation restrictions are left in place. It is important that policy relaxation be gradual, as this limits the spread of disease, and can allow for less restrictive measures in the future. In real-world terms, the gradual relaxation suggested by our model can correspond to the lifting of restrictions on in-person work operations several industries at a time, or to gradually increasing the number of low-risk people allowed to gather in one place. However, unless measures are relaxed slowly, a second outbreak occurs, so recovery of economic output is slow. Aside from a trade-off between health and economic outcomes, our model illustrates a trade-off between short-term and long-term outcomes which may not always be perceived.

These results also suggest that optimal policies may differ in various countries independent of the level of disease spread and healthcare capacity. For instance, in countries with more vulnerable economies that have a younger population, the consequences from disease may be less than in other countries, while consequences from isolation policies may be higher than in other countries. Countries with lower hospital capacity could face higher consequences from less restrictive isolation policies. Our model framework can easily be applied to the analysis of countries besides the United States. Our code is publicly available for further research.

We also see that contagiousness of non-hospitalized infected individuals (*β*) has an important effect on disease spread and mortality, and this suggests that strictly enforcing isolation measures is especially important for policymakers to prioritize. Measures that decrease spread out of isolation, such as universal mask-wearing, and increased testing to inform non-hospitalized infected individuals of their status could reduce contagiousness. Lowering *β* could allow for isolation measures to be relaxed more quickly. However, if policymakers prioritize short-term economic productivity more, their isolation policies may lead to many times more deaths and hospitalizations with minimal short-term economic gain.

Our results demonstrate that keeping stringently isolating at least a substantial portion of the population is necessary to avert large numbers of deaths. This is exemplified by the large number of deaths observed when the isolation measures are lifted rapidly.

More investigation is required as to how to keep such isolation measures sustainable over time, both through targeted relaxation, as we suggest, but also through safe provision of services to the isolated and increased infrastructure for at-home economic productivity (30). More workers may be required for maintaining an isolated population over time. Additionally, continued compliance with isolation restrictions may require a variety of measures, including enforcement, education from public health officials, and infrastructure for alternatives to traditional social and entertainment options.

Furthermore, policymakers should use other tools to supplement isolation policies, including contact tracing and other containment measures. Successful isolation policy can more quickly bring disease levels down to a level where more targeted tools can control future outbreaks. Our simulations suggest that isolation policy alone cannot end a pandemic without a level of enforcement that may not be realistic, and one that necessitates extreme economic costs. Thus, policymakers should quickly incentivize vaccines and other medical treatments. Our study has several limitations. We do not model how individuals react to lengthy restrictions related to infectious disease. We also do not model any individual-level differences in behavior or disease spread. This model could also be modified to incorporate the effects of testing and to have probability of infection depend on job type (31). Our model does not allow for any differences in disease spread between new and repeated contacts, which some models have incorporated (32; 33; 34). We also model isolation restrictions as completely eliminating the risk of infection. The less effective isolation policies are at lowering the risk of infection, the less attractive isolation policies become. Since testing policies can allow isolation restrictions to become more targeted, incorporating them can make the non-isolated state less costly for disease spread. Additionally, due to the newness of COVID-19, the exact disease parameters and the effects of seasonality are not precisely known (35; 36). Because of the uncertainty surrounding disease parameters, which change as the COVID-19 situation progresses, and because of the limitations of our model, our numbers should not be taken as literal predictions, but rather as illustrating the consequences of different policy approaches and individual behaviors. Our model has applications to different types of populations, as well as to future pandemics.

During a fatal infectious disease outbreak of lengthy duration, making policy decisions with longer term consequences in mind is essential. Our model provides a framework for making such decisions that takes into account differences within the population and disease changes that may occur over time. We provide evidence about what isolation policies may allow for minimal deaths while maximizing economic productivity. We find that once infection levels are somewhat controlled, very gradual relaxation of the restrictions on younger groups can minimize health consequences and economic damage.

## Conclusion

Our study demonstrates that removing isolation policies rapidly after controlling the outbreak but before eradicating the disease leads to a second outbreak and eventually necessitates re-enforcement of isolation policies. This prolongs the duration of the pandemic with worse outcomes in terms of the number of hospitalized individuals, number of deaths, and economic output.

The best outcomes in terms of managing the epidemic and reducing the damage to the economy are obtained in the last scenario. In this scenario, an extremely strict isolation policy for the senior population, combined with a very gradual removal of mandatory isolation for younger citizens after the outbreak is largely controlled, can lessen economic damage without catastrophic increases in the number of hospitalized and dead individuals. However, if isolation policy for the senior population is not strictly enforced, there can be severe consequences. No isolation restriction approach can produce a return to normality within 76 weeks. Isolation policies can only be used as a bridge to a global vaccination program.

Lastly, our sensitivity analysis shows that strict isolation policies and a strong reduction of contagiousness from the non-hospitalized infected individuals can reduce the total number of hospitalizations and the number of deaths as compared to loosely enforced measures.

## Data Availability

The source code will be uploaded upon acceptance for publication.

## Recommendations

From our observations we recommend that:

1. The maximum possible efforts should be implemented to accelerate the development of a vaccine for COVID-19.
2. Extremely strict isolation policies should be enforced for the senior population. These policies could include strict stay-at-home orders for seniors with specialized delivery systems, requiring extensive personal protective equipment (PPE) equipment for individuals interacting with seniors, or determining special hours for seniors to go outside.
3. The non-senior population should remain under isolation policies until the number of new daily infections is drastically reduced. Then, the non-senior population could leave isolation very gradually.

